# Automated Evaluation for Pericardial Effusion and Cardiac Tamponade with Echocardiographic Artificial Intelligence

**DOI:** 10.1101/2024.11.27.24318110

**Authors:** I-Min Chiu, Milos Vukadinovic, Yuki Sahashi, Paul P. Cheng, Chi-Yung Cheng, Susan Cheng, David Ouyang

## Abstract

**Background:** Timely and accurate detection of pericardial effusion and assessment cardiac tamponade remain challenging and highly operator dependent.

**Objectives:** Artificial intelligence has advanced many echocardiographic assessments, and we aimed to develop and validate a deep learning model to automate the assessment of pericardial effusion severity and cardiac tamponade from echocardiogram videos.

**Methods:** We developed a deep learning model (EchoNet-Pericardium) using temporal-spatial convolutional neural networks to automate pericardial effusion severity grading and tamponade detection from echocardiography videos. The model was trained using a retrospective dataset of 1,427,660 videos from 85,380 echocardiograms at Cedars-Sinai Medical Center (CSMC) to predict PE severity and cardiac tamponade across individual echocardiographic views and an ensemble approach combining predictions from five standard views. External validation was performed on 33,310 videos from 1,806 echocardiograms from Stanford Healthcare (SHC).

**Results:** In the held out CSMC test set, EchoNet-Pericardium achieved an AUC of 0.900 (95% CI: 0.884– 0.916) for detecting moderate or larger pericardial effusion, 0.942 (95% CI: 0.917–0.964) for large pericardial effusion, and 0.955 (95% CI: 0.939–0.968) for cardiac tamponade. In the SHC external validation cohort, the model achieved AUCs of 0.869 (95% CI: 0.794–0.933) for moderate or larger pericardial effusion, 0.959 (95% CI: 0.945–0.972) for large pericardial effusion, and 0.966 (95% CI: 0.906–0.995) for cardiac tamponade. Subgroup analysis demonstrated consistent performance across ages, sexes, left ventricular ejection fraction, and atrial fibrillation statuses.

**Conclusions:** Our deep learning-based framework accurately grades pericardial effusion severity and detects cardiac tamponade from echocardiograms, demonstrating consistent performance and generalizability across different cohorts. This automated tool has the potential to enhance clinical decision-making by reducing operator dependence and expediting diagnosis.

## Introduction

Cardiac tamponade, a life-threatening condition, occurs when excess fluid accumulates in the pericardial space, resulting in increased intrapericardial pressure, impairs cardiac filling, and reduces cardiac output. Without prompt recognition and intervention, this can lead to shock and circulatory collapse. Echocardiography is the gold standard for pericardial effusion detection due to its accessibility, portability, and comprehensive assessment of both anatomy and function^1–3^. However, despite increasing availability of ultrasound technology, the accurate assessment of pericardial effusion and cardiac tamponade still depends on expert image acquisition and interpretation. Distinguishing the severity of pericardial effusion and presence of tamponade can be challenging, with mild effusions sometimes mistaken for pericardial fat, and more critically, the severity of effusion does not necessarily correlate with the risk of tamponade^4^.

These challenges highlight the need for a reliable, automated method for detecting and assessing pericardial effusions that minimizes operator dependence and effectively assesses the risk of tamponade. Artificial intelligence (AI) has shown significant success in evaluating echocardiography tasks, including the assessment of left ventricular function, wall motion abnormalities, right ventricular function, and valvular disease^5–9^. AI presents a promising approach to automating complex image interpretation, improving the precision of measurements, and identify subtle cardiac phenotypes. While previous studies assessing pericardial effusions with deep learning have been limited by small training datasets and lack of eternal validation^10,11^, such that further work is warranted to apply AI to assessing pericardial effusion severity and predicting progression to cardiac tamponade.

In this study, we developed and evaluated performance of a deep learning pipeline in automating identification of PE and cardiac tamponade from standard transthoracic echocardiogram studies. We hypothesized that a deep learning approach can identify and assess PE severity on combinations of standard echocardiogram view videos with high-throughput automation.

## Methods

### Study Populations and Data Collection

In this study, we analyzed previously collected transthoracic echocardiogram studies from Cedars-Sinai Medical Center (CSMC) between September 12, 2011, and June 4, 2022. Echocardiogram videos were originally stored as DICOM videos from GE or Philips ultrasound machines. We pre-processed these videos to remove non-ultrasound sector information, extracted metadata, and converted them to AVI format^12^. Videos were stored as 112 × 112-pixel video files and view classified into five standard echocardiographic views (apical-4-chamber, apical-2-chamber, parasternal long axis, parasternal short axis, and subcostal views). The view classifier model^13^ used in this study was trained on 77,426 echocardiogram videos to classify 58 specific view categories. Pericardial effusion severity and presence of cardiac tamponade for was extracted from the clinical report determined in a high-volume echocardiography laboratory in accordance with American Society of Echocardiography guidelines^14^. The model was trained on 4 classes of pericardial effusion sizes (none, mild, moderate, and large), with intermediate categories were assigned to the more severe category.

EchoNet-Pericardium - a full end-to-end approach including video processing, view classification, pericardial effusion size assessment, and identification of cardiac tamponade - was externally validated using data from an independent and geographically distinct high-volume echocardiography lab. The model was evaluated on 1,806 studies (containing a total of 33,310 videos) from the Stanford Healthcare (SHC). The ensemble prediction utilized predictions from five standard echocardiographic views for evaluating pericardial effusion size and tamponade model validation. Model output was compared with clinical grading determined by expert cardiologists from the clinical reports. This study was approved by the institutional review boards at CSMC and SHC.

### AI Model

Echocardiogram videos were divided into training, validation, and test datasets in an 8:1:1 ratio by patient, ensuring no patient overlap across the sets to prevent data leakage. If a patient had multiple echocardiogram studies or videos, each video was treated as an independent example during model training. Deep learning models were trained using the PyTorch Lightning deep learning framework. Video-based convolutional neural networks (R2+1D) were used for PE severity assessment and cardiac tamponade prediction^15^. This model architecture was previously used for other echocardiography tasks and shown to be effective^6,9^. The models were initialized with random weights and trained using a binary cross entropy loss function for up to 50 epochs, using an ADAM optimizer, an initial learning rate of 1e-2, and a batch size of 64 on NVIDIA RTX A6000 GPU. Early stopping was performed based on the validation loss if no improvement after 5 epochs. For model training, we selected random clips of 32 frames, sampled every other frame to obtain 16 frames per clip, and input these into the model. For pericardial effusion size assessment, models were trained for each individual view, with input videos of each individual view, and an ensemble model was constructed by logistical regression with inputs of the inference prediction from models of each view. For tamponade prediction, we trained the model using all apical-4-chamber videos for binary prediction (Central Illustration).

### Statistical Analysis

Continuous variables are expressed as mean ± standard deviation, while nominal variables are presented as proportions. All analyses were performed on the held-out test dataset and external test set, which was never seen during model training. Model performance was evaluated using area under the receiver operating characteristic curve and confusion matrices. Confidence intervals were computed using 1,000 bootstrapped samples of the test datasets. Statistical analysis was performed in Python (version 3.8.0). Subgroup analysis was conducted to assess model performance in patients with different age, sex, race, and other clinical characteristics. Clinical characteristics were obtained from the electronic health record or associated echocardiography report.

## Results

### Primary cohort characteristics

A total of 85,380 transthoracic echocardiograms from 49,598 patients at CSMC, collected between September 12, 2011, and June 4, 2022, were used to train and evaluate the deep learning models. The mean (SD) age of the patients was 68.7 (19.4) years, with 49.7% being male. Among them, 3.5% had atrial fibrillation, 12.2% had heart failure, 44.9% had hypertension, and 19.4% had diabetes mellitus. Across all echocardiograms, 14.0% showed small pericardial effusion, 4.2% had moderate pericardial effusion, 1.1% had large pericardial effusion, and 0.7% presented with cardiac tamponade (Table 1). Echocardiograms were divided into training, validation, and test datasets in an 8:1:1 ratio by patient, resulting in 68,110 echocardiograms in the training set, 8,508 in the validation set, and 8,762 in the test set. A total of 1,427,660 videos were included through the view classifier process, with 363,817 from A4C, 179,361 from A2C, 355,330 from PLAX, 294,950 from PSAX, and 234,202 from Subcostal view (Supplemental Table 1).

**Table 1.** Patient Characteristics.

| Characteristic | CSMC |  |  | SHC |
| --- | --- | --- | --- | --- |
|  | Train | Validation | Test |  |
| Number of patients | 39,654 | 4,934 | 5,010 | 1,806 |
| Sex (% Male) | 19,711 (49.7) | 2,438 (49.4) | 24,90 (49.7) | 957 (53.0) |
| Age, mean (std), y | 68.7 (19.4) | 68.5 (19.3) | 68.6 (19.3) | 60.4 (17.3) |
| Race |  |  |  |  |
| Caucasian, n (%) | 26,944 (67.9%) | 3,393 (68.8%) | 3,419 (68.2%) | 907 (50.2) |
| Black, n (%) | 5,774 (14.6%) | 717 (14.5%) | 734 (14.7%) | 76 (4.2) |
| Asian, n (%) | 2,981 (7.5%) | 343 (7.0%) | 364 (7.3%) | 303 (16.8) |
| Others, n (%) | 3,955 (10.0%) | 481 (9.7%) | 493 (9.8%) | 163 (9.0) |
| Clinical History |  |  |  |  |
| Atrial Fibrillation | 1,375 (3.5%) | 179 (3.6%) | 190 (3.8%) | 542 (30.0) |
| Heart Failure | 4,828 (12.2%) | 616 (12.5%) | 602 (12.0%) | 751 (41.6) |
| CAD | 8,796 (22.2%) | 1,138 (23.1%) | 1,094 (21.8%) | 298 (16.5) |
| Hypertension | 17,811 (44.9%) | 2,221 (45.0%) | 22,62 (45.1%) | 1,082 (59.9) |
| Diabetes Mellitus | 7,679 (19.4%) | 994 (20.1%) | 954 (19.0%) | 168 (9.3) |
| Number of Study | 68,110 | 8,508 | 8,762 | 1,806 |
| Number of Video | 1,139,533 | 142,341 | 145,786 | 33,310 |
| Pericardial Effusion |  |  |  |  |
| Small | 9,512 (14.0%) | 1,193 (13.6%) | 1,242 (14.2%) | 83 (4.6%) |
| Moderate | 2,831 (4.2%) | 336 (3.9%) | 383 (4.4%) | 19 (1.1%) |
| Large | 796 (1.2%) | 77 (0.9%) | 78 (0.9%) | 11 (0.6%) |
| Cardiac Tamponade | 482 (0.7%) | 64 (0.8%) | 39 (0.4%) | 17 (0.9%) |

### Model performance in the primary cohort

We trained five view-specific deep learning models and ensembled them to obtain the final pericardial effusion size assessment (Table 2). Our ensemble model effectively distinguished PE severity, achieving an AUC of 0.900 (0.884-0.916) for detecting moderate or above PE and 0.942 (0.917-0.964) for large PE in the primary cohort (Figure 1A). For cardiac tamponade, the model, trained using A4C views, achieved an AUC of 0.955 (0.939 - 0.968). When evaluated on echocardiogram studies with pericardial effusion, the model distinguished cases of tamponade with an AUC of 0.904 (0.881 - 0.924) (Figure 2A).

**Figure 1.**
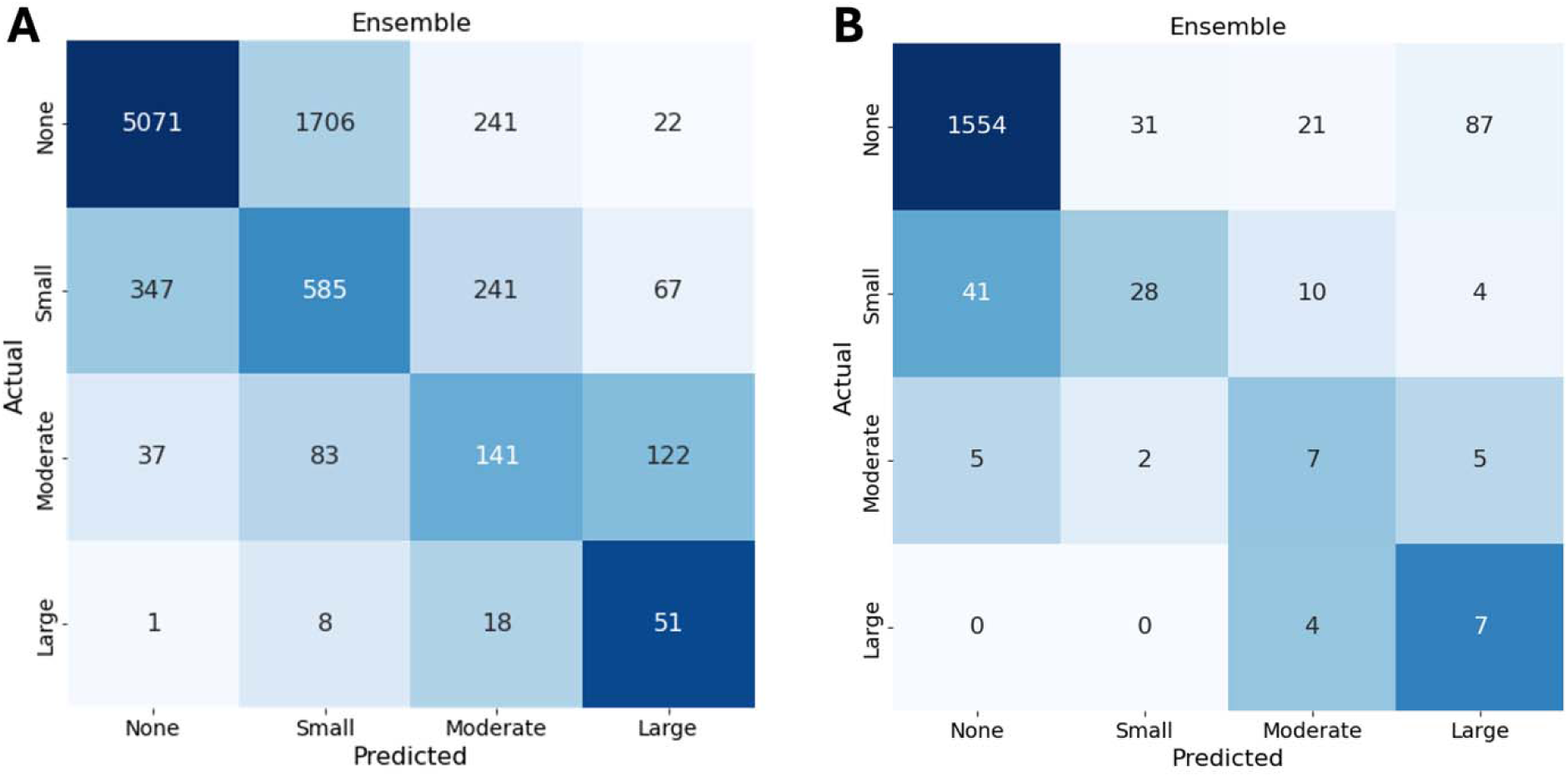
Confusion matrices for pericardial effusion severity classification in the primary cohort (A) and the external validation cohort (B). The EchoNet-Pericardium model utilizes an ensemble of predictions from five standard echocardiographic views to classify pericardial effusion as none, small, moderate, or large. Figure 1A demonstrates performance in the primary test cohort, while 1B depicts results in the external validation cohort. The diagonal values indicate correct predictions, with the model showing consistent accuracy across different levels of effusion severity.

**Figure 2.**
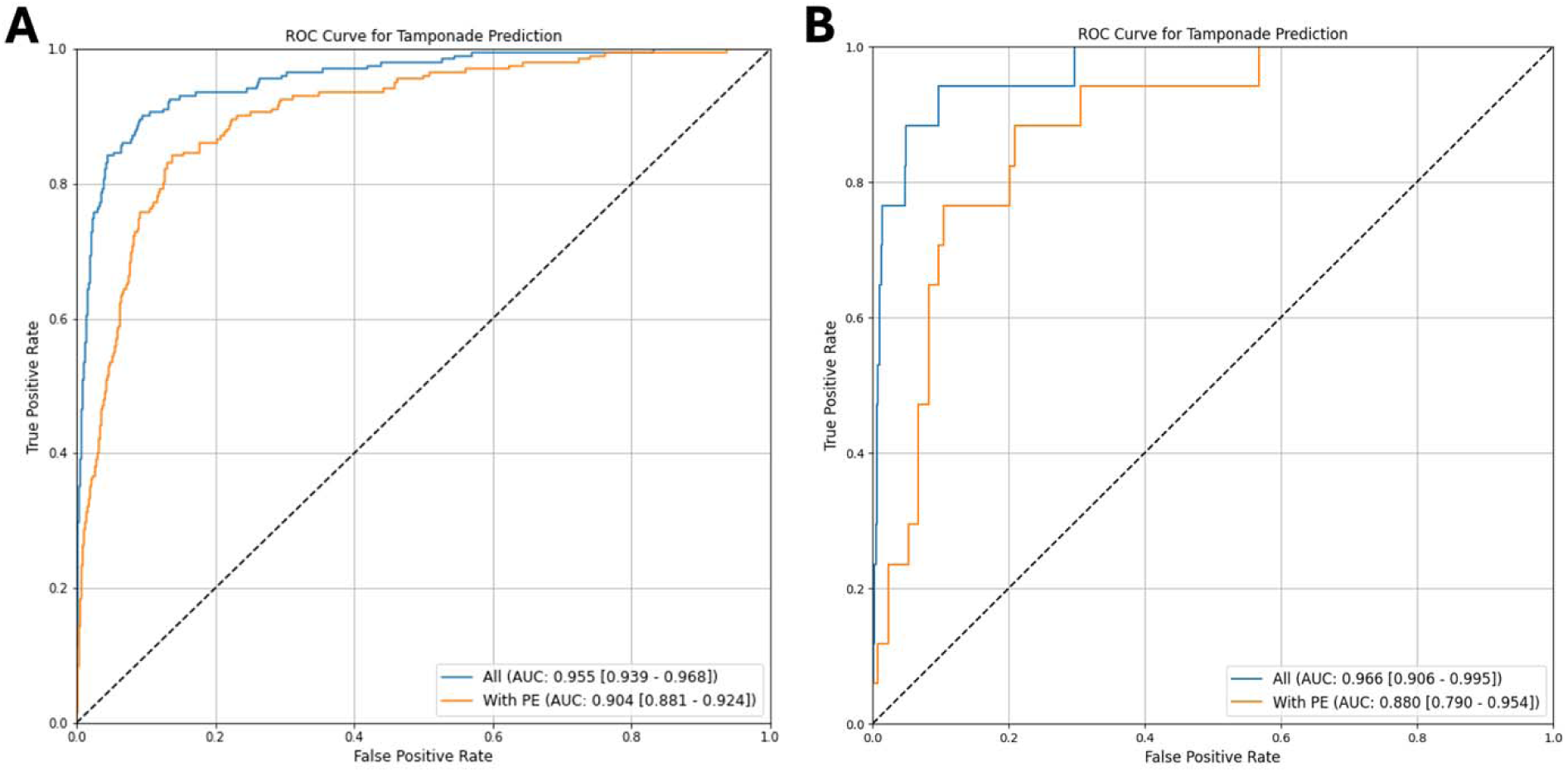
Receiver operating characteristic curves for cardiac tamponade prediction in the primary cohort (A) and the external validation cohort (B). The EchoNet-Pericardium model distinguishes cardiac tamponade cases using apical-4-chamber (A4C) views. The blue curves represent model performance on all echocardiograms, while the orange curves show performance specifically for studies with pericardial effusion. Figure 2A corresponds to the primary test cohort, achieving an AUC of 0.955 for all echocardiograms and 0.904 for cases with effusion. Figure 2B represents the external validation cohort, with an AUC of 0.966 for all echocardiograms and 0.880 for cases with effusion.

**Table 2.** EchoNet-Pericardium Model Performance.

| Characteristic | Small PE | Moderate PE | Large PE |
| --- | --- | --- | --- |
| CSMC |  |  |  |
| Apical 4 Chamber | 0.779 (0.766 - 0.792) | 0.890 (0.875 - 0.904) | 0.944 (0.918 - 0.965) |
| Apical 2 Chamber | 0.734 (0.720 - 0.747) | 0.841 (0.818 - 0.862) | 0.872 (0.815 - 0.926) |
| Parasternal long axis | 0.645 (0.633 - 0.658) | 0.745 (0.721 - 0.770) | 0.837 (0.785 - 0.880) |
| Parasternal short axis | 0.710 (0.697 - 0.723) | 0.810 (0.788 - 0.832) | 0.887 (0.842 - 0.924) |
| Subcostal | 0.630 (0.617 - 0.643) | 0.722 (0.696 - 0.747) | 0.761 (0.701 - 0.819) |
| Ensemble of all views | 0.805 (0.792-0.816) | 0.900 (0.884-0.916) | 0.942 (0.917-0.964) |
| SHC |  |  |  |
| Ensemble of all views | 0.746 (0.697 - 0.790) | 0.869 (0.794 - 0.933) | 0.959 (0.945 - 0.972) |

As shown in the confusion matrix, the model’s performance in the test set of the primary cohort for detecting moderate or larger pericardial effusion demonstrated a sensitivity of 0.72 (0.698 – 0.741), a specificity of 0.931 (0.913 – 0.949), and a negative predictive value (NPV) of 0.984 (0.969 – 0.999). For detecting large pericardial effusions, the model achieved a sensitivity of 0.654 (0.622 – 0.686), a specificity of 0.976 (0.958 – 0.994), and an NPV of 0.997 (0.981 – 1.0). These metrics indicate the model’s accuracy in identifying and excluding cases of moderate or severe PE and large PE.

### Subgroup Analysis

In the subgroup analysis, the model demonstrated consistent performance in predicting moderate or larger pericardial effusion and cardiac tamponade across various subgroups within the test set (Table 3). For patients over 65 years, the AUC for moderate or larger pericardial effusion was 0.901 (95% CI, 0.882–0.921), and for tamponade, it was 0.953 (95% CI, 0.934–0.970). Performance across gender was similar, with an AUC of 0.896 (95% CI, 0.872–0.918) in males and 0.902 (95% CI, 0.881–0.923) in females for moderate or larger pericardial effusion, and 0.974 (95% CI, 0.960–0.984) in males and 0.944 (95% CI, 0.921–0.964) in females for tamponade.

**Table 3.** Subgroup Analysis.

| Characteristic | Case | Moderate PE or worse | Tamponade |
| --- | --- | --- | --- |
| All | 8762 | 0.900 (0.884-0.916) | 0.955 (0.939 - 0.969) |
| Age > 65 years old | 5319 | 0.901 (0.882 - 0.921) | 0.953 (0.934 - 0.970) |
| Age < 65 years old | 3443 | 0.897 (0.868 - 0.922) | 0.955 (0.925 - 0.977) |
| Male | 4388 | 0.896 (0.872 - 0.918) | 0.974 (0.960 - 0.984) |
| Female | 4374 | 0.902 (0.881 - 0.923) | 0.944 (0.921 - 0.964) |
| Caucasian | 5955 | 0.891 (0.870 - 0.911) | 0.959 (0.942 - 0.972) |
| Black | 1323 | 0.899 (0.864 - 0.931) | 0.936 (0.892 - 0.975) |
| Asian | 668 | 0.951 (0.925 - 0.972) | -- |
| LVEF > 50 | 6875 | 0.908 (0.891 - 0.925) | 0.970 (0.956 - 0.982) |
| LVEF < 50 | 1688 | 0.898 (0.855 - 0.930) | 0.900 (0.848 - 0.942) |
| Atrial fibrillation during examination | 378 | 0.901 (0.831 - 0.951) | 0.979 (0.969 - 0.988) |
| Without Atrial fibrillation during examination | 8384 | 0.900 (0.882 - 0.915) | 0.955 (0.939 - 0.970) |

The model’s performance was further evaluated across different ranges of left ventricular ejection fraction (LVEF). In patients with LVEF below 50, the model achieved AUCs of 0.898 (95% CI, 0.855–0.930) for moderate or larger pericardial effusion and 0.899 (95% CI, 0.848–0.942) for tamponade. Among patients with atrial fibrillation during examination, the model achieved an AUC of 0.901 (95% CI, 0.831–0.951) for moderate or larger pericardial effusion and 0.979 (95% CI, 0.969–0.988) for tamponade, while in patients without atrial fibrillation, the AUCs were 0.900 (95% CI, 0.882–0.915) and 0.955 (95% CI, 0.939–0.970) for moderate or larger pericardial effusion and tamponade, respectively.

### External validation

The external validation cohort from SHC consisted of 1,806 patients with a mean (SD) age of 60.4 (17.3) years, and 53.0% of them were male. Regarding race, 50.2% of the patients were Caucasian, 4.2% were Black, and 16.8% were Asian. Clinical history data revealed that 30.0% of the patients had atrial fibrillation, 41.6% had heart failure, 16.5% had CAD, 59.9% had hypertension, and 9.3% had diabetes mellitus. The dataset included 1,806 echocardiograms, corresponding to 33,310 videos. Among these, 4.6% showed small pericardial effusion, 1.1% had moderate pericardial effusion, 0.6% had large pericardial effusion, and 0.9% of cases presented with cardiac tamponade. A total of 33,310 videos were included through the view classifier process, with 9,462 from A4C, 4,040 from A2C, 7,982 from PLAX, 6,601 from PSAX, and 5,225 from Subcostal view (Supplemental Table 2).

The EchoNet-Pericardium showed consistent performance in distinguishing pericardial effusion size and tamponade (Figure 1B) in the external validation cohort. The model demonstrated an AUC of 0.869 (0.794 - 0.933) in detecting at least moderate pericardial effusion and an AUC of 0.959 (0.945 - 0.972) for large pericardial effusion. For cardiac tamponade, the model achieved an AUC of 0.966 (0.906 - 0.995). When evaluated on echocardiograms with at least a small pericardial effusion, the model distinguished cases of tamponade with an AUC of 0.880 (0.790 - 0.954) (Figure 2B). As shown in the confusion matrix, the model’s performance in the external validation cohort for detecting moderate or larger pericardial effusion demonstrated a sensitivity of 0.737 (0.539 – 0.935), a specificity of 0.976 (0.968 - 0.983), and an NPV of 0.997 (0.994 - 1.0). For detecting large pericardial effusions, the model achieved a sensitivity of 0.636 (0.352 - 0.921), a specificity of 0.943 (0.932 - 0.954), and an NPV of 0.997 (0.995 - 1.0).

## Discussion

This study presents a validated deep learning framework capable of accurately assessing PE severity and detecting cardiac tamponade using echocardiography. From a full transthoracic echocardiogram, EchoNet-Pericardium effectively identified pericardial effusion size from five standard echocardiographic views, achieving an AUC of 0.900 for moderate or larger pericardial effusion and 0.942 for large PE in primary cohorts. Our model also demonstrated high accuracy in detecting cardiac tamponade with an AUC of 0.955. EchoNet-Pericardium demonstrated consistent performance across a geographically distinct external validation cohort without preselection, showcasing its robustness and generalizability in real-world settings. Implementing this automated tool could expedite diagnosis, reduce operator dependence, and potentially improve patient outcomes by enabling timely interventions.

For a comprehensive clinical evaluation of pericardial effusions and cardiac tamponade, clinicians utilize a variety of echocardiographic views are assessed together. In EchoNet-Pericardium, the use of video-based convolutional neural networks (R2+1D) trained for each view and then ensembled captures temporal dynamics crucial for identifying pericardial effusion size and distinguishing cases with tamponade. For classifying tamponade, we chose the A4C view to focus on given the appearance of all four chambers that allows the model to assess intraventricular dependence and associations between the left and right heart. Our study builds on the intersection of AI and echocardiography, advancing previous achievements in cardiovascular imaging applications. Significant progress has been made in automating view classification^16,17^, identifying structural abnormalities like left ventricular hypertrophy^8,18^, evaluating left ventricular systolic function^6^, and assessing valvular function and vascular emergency^9,19,20^. By integrating our model with existing AI-driven innovations, we aim to support less experienced operators in obtaining high-quality imaging^21,22^, thereby improving diagnostic accuracy for pericardial effusion and enabling early detection of tamponade. Future research could focus on refining automated quantification, validating model performance across varied clinical settings, and expanding access to diagnostic tools in primary care and resource-limited regions.

This study has certain limitations that should be considered when interpreting the results. First, although the model was trained on a large-scale dataset, the retrospective nature of our data collection may limit its performance in real-time clinical settings. Second, while we included diverse echocardiographic views to improve the accuracy of pericardial effusion grading, this model has yet to be tested in point-of-care ultrasonography performed by noncardiologists. Further research is needed to assess the model’s performance with videos of varying quality and levels of operator expertise. Although our analyses across different health systems indicate that the deep learning algorithm is robust to variations in practice patterns across regions, additional work and prospective validation are necessary to better understand the impact of AI-guided screening workflows on clinical care.

In conclusion, our study presents a novel, automated approach for detecting and grading pericardial effusion severity and identifying cardiac tamponade using deep learning models applied to echocardiography. EchoNet-Pericardium’s consistent performance across multiple datasets highlights its potential to improve clinical decision-making and streamline echocardiographic assessment of PE and cardiac tamponade. Prospective studies and integration into clinical practice will be essential to fully realize the benefits of this technology.

## Data availability

The dataset of videos and reports used to train EchoNet-Pericardium is not publicly available due to its potentially identifiable nature.

## Code Availability

The code for EchoNet-Pericardium will be made publicly available following the publication of the paper.

---

**Central Illustration.**
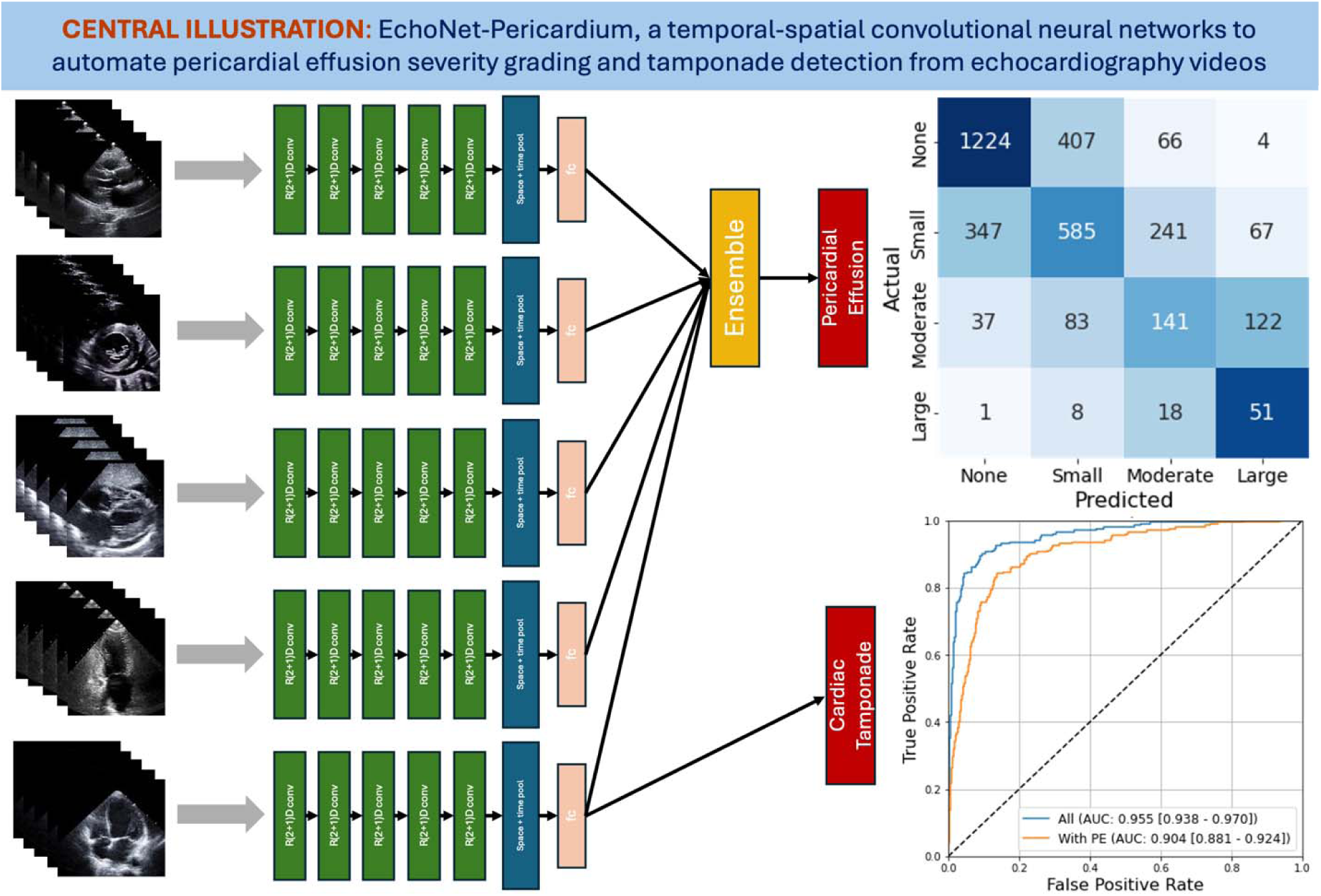
EchoNet-Pericardium: A temporal-spatial convolutional neural network designed to automate pericardial effusion severity grading and tamponade detection using echocardiography videos. The model integrates predictions from five standard echocardiographic views through an ensemble approach, achieving an AUC of 0.900 for detecting moderate or larger pericardial effusion, 0.942 for large pericardial effusion, and 0.955 for cardiac tamponade.

