## Supplemental Table for "Automated Evaluation for Pericardial Effusion and Cardiac Tamponade with Echocardiographic Artificial Intelligence"

Supplemental Table 1. Cedars Sinai Medical Center Patient Characteristics

|  | CSMC | | | | |
| --- | --- | --- | --- | --- | --- |
| Characteristic | A4C | A2C | PLAX | PSAX | Subcostal |
| Number of patients | 49,030 | 46,782 | 49,223 | 48,746 | 47,567 |
| Sex (% Male) | 24,336 (49.6) | 23,285 (49.8) | 24,426 (49.6) | 24,108 (49.5) | 23,490 (49.4) |
| Age, mean (std), y | 68.2 (18.3) | 68.2 (18.2) | 68.2 (18.3) | 68.2 (18.3) | 68.2 (18.2) |
| Race |  |  |  |  |  |
| Caucasian, n (%) | 33,345 (68.0%) | 31,818 (68.0%) | 33,506 (68.1%) | 33,145 (68.0%) | 32,359 (68.0%) |
| Black, n (%) | 7,160 (14.6%) | 6,803 (14.5%) | 7,167 (14.6%) | 7,121 (14.6%) | 6957 (14.6%) |
| Asian, n (%) | 3,657 (7.5%) | 3,516 (7.5%) | 3,665 (7.4%) | 3,639 (7.5%) | 3,560 (7.5%) |
| Others, n (%) | 4,868 (9.9%) | 4,645 (9.9%) | 4,885 (9.9%) | 4,841 (9.9%) | 4,691 (9.9%) |
| Clinical History |  |  |  |  |  |
| Atrial Fibrillation | 1,731 (3.5%) | 1,647 (3.5%) | 1,728 (3.5%) | 1,718 (3.5%) | 1,665 (3.5%) |
| Heart Failure | 5,991 (12.2%) | 5,737 (12.3%) | 6,005 (12.2%) | 5,949 (12.2%) | 5,783 (12.2%) |
| CAD | 10,913 (22.3%) | 10,423 (22.3%) | 10,946 (22.2%) | 10,832 (22.2%) | 10,557 (22.2%) |
| Hypertension | 22,064 (45.0%) | 21,106 (45.1%) | 22,134 (45.0%) | 21,913 (45.0%) | 21,407 (45.0%) |
| Diabetes Mellitus | 9,530 (19.4%) | 9,118 (19.5%) | 9,552 (19.4%) | 9,450 (19.4%) | 9,214 (19.4%) |
| Number of Studies | 83,824 | 77,427 | 84,408 | 83,057 | 80,072 |
| Number of Videos | 363,817 | 179,361 | 355,330 | 294,950 | 234,202 |
| Pericardial Effusion |  |  |  |  |  |
| Small | 11,701 (14.0%) | 10,520 (13.6%) | 11,820 (14.0%) | 11,626 (14.0%) | 11,179 (14.0%) |
| Moderate | 3,460 (4.1%) | 2,917 (3.8%) | 3,521 (4.2%) | 3453 (4.2%) | 3,411 (4.3%) |
| Large | 928 (1.1%) | 671 (0.9%) | 934 (1.1%) | 912 (1.1%) | 915 (1.1%) |
| Cardiac Tamponade | 566 (0.7%) | 406 (0.5%) | 578 (0.7%) | 552 (0.7%) | 570 (0.7%) |

*A4C: Apical 4 chamber, A2C: Apical 2 chamber, PLAX: parasternal long axis, PSAX: parasternal short axis, CAD: coronary artery disease

Supplemental Table 2. Stanford HealthCare Patient Characteristics

|  | SHC | | | | |
| --- | --- | --- | --- | --- | --- |
| Characteristic | A4C | A2C | PLAX | PSAX | Subcostal |
| Number of patients | 1,804 | 1,745 | 1,756 | 1,750 | 1,739 |
| Sex (% Male) | 971 (53.8) | 941 (53.9) | 945 (53.8) | 942 (53.8) | 937 (53.9) |
| Age, mean (std), y | 59.7 (17.2) | 59.8 (17.3) | 59.7 (17.2) | 59.7 (17.3) | 59.7 (17.3) |
| Race |  |  |  |  |  |
| Caucasian, n (%) | 958 (53.1%) | 930 (53.3%) | 935 (53.2%) | 934 (53.4%) | 928 (53.4%) |
| Black, n (%) | 80 (4.4%) | 75 (4.3%) | 78 (4.4%) | 77 (4.4%) | 73 (4.2%) |
| Asian, n (%) | 295 (16.3%) | 290 (16.6%) | 293 (16.7%) | 292 (16.7%) | 287 (16.5%) |
| Others, n (%) | 472 (26.1%) | 450 (25.8%) | 450 (25.6%) | 447 (25.5%) | 451 (25.9%) |
| Clinical History |  |  |  |  |  |
| Atrial Fibrillation | 369 (20.5%) | 354 (20.3%) | 358 (20.4%) | 359 (20.5%) | 359 (20.6%) |
| Heart Failure | 526 (29.2%) | 510 (29.3%) | 515 (29.3%) | 515 (29.4%) | 515 (29.6%) |
| CAD | 448 (24.8%) | 438 (25.1%) | 435 (24.8%) | 436 (24.9%) | 435 (25.0%) |
| Hypertension | 905 (50.2%) | 880 (50.4%) | 887 (50.2%) | 880 (50.3%) | 877 (50.4%) |
| Diabetes Mellitus | 434 (24.1%) | 424 (24.3%) | 423 (24.1%) | 425 (24.3%) | 425 (24.4%) |
| Number of Studies | 1,804 | 1,745 | 1,756 | 1,750 | 1,739 |
| Number of Videos | 9,462 | 4,040 | 7,982 | 6,601 | 5,225 |
| Pericardial Effusion |  |  |  |  |  |
| Small | 113 (6.3%) | 108 (6.2%) | 109 (6.2%) | 108 (6.2%) | 106 (6.1%) |
| Moderate | 30 (1.7%) | 29 (1.7%) | 29 (1.7%) | 29 (1.7%) | 27 (1.6%) |
| Large | 11 (0.6%) | 9 (0.5%) | 11 (0.6%) | 9 (0.5%) | 9 (0.5%) |
| Cardiac Tamponade | 24 (1.3%) | 16 (0.9%) | 17 (1.0%) | 15 (0.9%) | 17 (1.0%) |

*A4C: Apical 4 chamber, A2C: Apical 2 chamber, PLAX: parasternal long axis, PSAX: parasternal short axis, CAD: coronary artery disease
